# Shifting Prevalence and Risk Factors of Non-communicable Diseases in Bangladesh: A Comparative Multilevel Analysis of Nationally Representative BDHS Data (2017–2022)

**DOI:** 10.64898/2026.03.31.26349897

**Authors:** Kazi Sabbir Ahmad Nahin, Arman Hossen, Tabita Jannatul

## Abstract

**Background:** Non-communicable diseases (NCDs) are significant public health concerns in Bangladesh, placing a heavy burden on the healthcare system. While the situation before COVID-19 was well-documented, it is unclear how the pandemic has impacted the prevalence and risk factors of these diseases. This study provides the first comparative assessment of the prevalence and determinants of diabetes mellitus (DM) and hypertension (HTN) before and after the pandemic, utilizing comprehensive multilevel data source and mixed-effects modeling to capture the shifting epidemiological burden.

**Methods:** We analyzed biomarker data from two nationally representative Bangladesh Demographic and Health Surveys (BDHS) 2017-18 and 2022. Diagnosis followed WHO guidelines for fasting blood glucose and blood pressure. Mixed-effect logistic regression models were employed to identify risk factors while accounting for the hierarchical survey design. The Intra-class Correlation Coefficient (ICC) was calculated to quantify the proportion of variance attributable to unobserved community-level heterogeneity.

**Results:** The study indicates a profound shift in the national burden of NCDs. Diabetes prevalence more than doubled, from 23% in 2017-18 to 49% in 2022, while hypertension prevalence declined from 22% to 15%, a pattern that may reflect survival bias among individuals with severe comorbidities. The previously strong bidirectional association between DM and HTN weakened in the post-pandemic period, hypertension continued to predict diabetes (AOR = 1.17), but diabetes was no longer a significant predictor of hypertension. Community-level determinants became substantially more influential, with local environmental factors playing a much larger role in shaping diabetes prevalence compared to the pre-pandemic period. Urban residence emerged as a significant new risk factor for diabetes in 2022 (AOR = 1.62; 95% CI: 1.34-1.96). Furthermore, the socioeconomic gap in diabetes risk narrowed as the disease affected more wealth groups, while higher educational attainment continued to serve as a protective factor against hypertension (AOR = 0.64; 95% CI: 0.54-0.75).

**Conclusion:** The post-pandemic landscape of NCDs in Bangladesh shows a clear divergence, marked by a rapid increase in diabetes contrasted with a stabilization in hypertension prevalence. Through comparative mixed-effects modeling, this study advances beyond simple prevalence comparisons to demonstrate the growing impact of urban environments and community-level factors on metabolic health. These evolving patterns underscore the need for integrated public health strategies that address emerging environmental risks and geographically specific vulnerabilities to support progress toward Sustainable Development Goal Target□3.4.

## Background

Non-communicable diseases (NCDs) account for 71% of global fatalities, resulting in the death of 41 million individuals annually—more than all other causes combined (1). In 2019, non-communicable diseases (NCDs) represented 74% of global fatalities and constituted 7 of the 10 principal causes of death (2). Disturbingly, 15 million individuals aged 30-69 succumb to premature mortality from non-communicable diseases (NCDs) each year, with 85% of these fatalities occurring in low- and middle-income countries (LMICs) (1).

Non-communicable diseases (NCDs) impact people at every stage of life, from infancy through old age, creating severe risks for public health, economic resilience, and social progress (3). Recent findings from the Lancet Taskforce on NCDs and Economics underscore the critical link between economic growth and disease management, stressing how poverty significantly worsens the impact of NCDs (4–8). The financial weight these diseases place on healthcare systems and national well-being is already heavy and is predicted to escalate in the near future (9).

Estimates suggest that by 2030, deaths linked to NCDs will jump from 41 million to 52 million, a trend that jeopardizes the success of Sustainable Development Goal (SDG) target 3.4, which seeks to cut premature NCD mortality by one-third through better prevention and care (10–12). Although wealthier nations have seen some success in managing these conditions, low- and middle-income countries (LMICs) continue to face steep challenges driven by persistent risk factors.

The World Health Organization (WHO) highlights four primary global drivers of NCDs: tobacco consumption, physical inactivity, alcohol abuse, and poor nutrition (13). These behaviors trigger four specific metabolic disruptions: high blood pressure (hypertension), obesity, high blood sugar (hyperglycemia), and abnormal lipid levels (hyperlipidemia) (1, 14).

Diabetes mellitus (DM) stands out as a major cause of death among NCDs, claiming a life every five seconds (15). As of 2017, roughly 451 million adults aged 18-99 were living with the condition, a figure expected to reach 693 million by 2045 (16). It is particularly concerning that nearly half—49.7%—of those with diabetes remain undiagnosed (17). LMICs shoulder 80% of this global burden, where cases often go undetected for years (18). In 2019 alone, diabetes was the direct cause of 1.5 million deaths, making it the ninth leading killer worldwide (19). South Asia faces a looming crisis, with prevalence rates expected to surge by more than 150% by 2030 (20).

Similarly, Hypertension (HTN), frequently called the ‘silent killer,’ is a primary driver of early death globally (21). It claims 9.4 million lives annually, a toll that rivals the mortality burden of major infectious diseases (22). By 2025, the number of people affected globally is expected to rise by 60%, reaching 1.56 billion (23). Despite common misconceptions, two-thirds of the world’s hypertension cases actually occur in LMICs (21, 24). In South Asia, including the SAARC nations, high blood pressure rates are already exceeding the global average (25).

The onset of NCDs is driven by a complicated mix of socioeconomic, demographic, and lifestyle choices (26–31). One major concern across both urban and rural populations is rising BMI, which is closely tied to higher diabetes risk (30–32). Interestingly, recent data points to an ‘inverted U-shaped’ relationship between socioeconomic status (SES) and the rates of DM and HTN (33). Smoking acts as another key risk factor for Type 2 Diabetes, particularly for heavy smokers. This elevated risk persists for up to a decade after quitting, though it drops faster for those who smoked less (34–36). Finally, while gender is consistently viewed as a significant factor, exposure to media seems to have extraordinarily little impact on the prevalence of these diseases (37).

As an LMIC, Bangladesh is grappling with a severe burden of NCDs, which now account for 59% of all mortality (38, 39). A systematic review and meta-analysis placed the national prevalence of diabetes at approximately 7.8% (95% CI: 6.4-9.3) (40). World Health Organization (WHO) data indicates that 12.88 million people, about 8% of the population live with diabetes in Bangladesh, a condition responsible for 3% of total deaths (41). Hypertension is rapidly becoming a major concern as well, with recent findings showing a prevalence rate of 20% across the country (21, 42).

The burden of Type 2 diabetes mellitus (T2DM) is notably rising in both urban centers and rural communities (43). Although religious norms keep alcohol consumption low, other behavioral risks—specifically tobacco use, sedentary lifestyles, and poor dietary habits—are trending upward (14, 44). Geographic analyses highlight that factors like age, education, and socioeconomic status (SES) are primary drivers of HTN and T2DM, often leading to significant regional inequalities (45, 46). Current literature has pinpointed a cluster of covariates linked to the high rates of DM and HTN in Bangladesh; these include gender, advanced age, higher educational attainment, wealth, elevated BMI, unemployment, urban living, and smoking (47–50).

The escalating prevalence of DM and HTN continue to place an immense burden on global healthcare systems and well-being (15, 51–54). In Bangladesh, the World Health Organization (WHO) has collaborated with the government to implement a multisectoral action plan for NCD prevention involving diverse ministries. However, identifying static risk factors is no longer sufficient; understanding the trajectory of these diseases is critical for effective resource allocation (55).

While previous studies have utilized the Bangladesh Demographic and Health Survey (BDHS) 2017-18 to establish a pre-pandemic baseline (56), there is a critical knowledge gap regarding how the prevalence and determinants of NCDs have shifted in the post-pandemic landscape. The interval between the BDHS 2017-18 and the newly released BDHS 2022 encompasses the COVID-19 pandemic, a period characterized by lockdowns, restricted healthcare access, increased psychological stress, and potential shifts in sedentary behavior, all of which are potent drivers of NCD risk.

Furthermore, the COVID-19 pandemic had direct impacts on the healthcare system and the human body. Severe interruptions to regular healthcare services meant that many people could not access routine screening and management for chronic diseases, which likely increased the number of undiagnosed cases (57). At the same time, clinical evidence shows that the SARS-CoV-2 infection itself can cause severe inflammation in the body. This inflammation can damage blood vessels and might accelerate the development of conditions like diabetes (58). Because of all these combined behavioral, healthcare, and biological changes, it is necessary to evaluate how the vulnerabilities for NCDs at both the individual and community levels have changed during this crisis.

Consequently, this study aims to conduct a comparative analysis of the sociodemographic, socioeconomic, and lifestyle-related determinants of DM and HTN using two nationally representative datasets: BDHS 2017-18 and BDHS 2022. By quantifying the temporal changes in disease prevalence and risk factors, we aim to provide policymakers with evidence-based insights into the pandemic’s ‘shock’ effect on NCD trends. This analysis is vital for recalibrating public health interventions to mitigate the compounding financial and social strain of NCDs and is essential for tracking Bangladesh’s progress toward achieving Sustainable Development Goal (SDG) Target 3.4 (reducing premature mortality from NCDs by one-third) in a post-COVID era.

## Methods

### Data Source and Study Design

This study utilizes data from two successive rounds of the Bangladesh Demographic and Health Survey (BDHS): the 2017-18 and 2022 iterations (59, 60). Both surveys were implemented by the National Institute of Population Research and Training (NIPORT) under the Ministry of Health and Family Welfare, as part of the global DHS program. These surveys provide nationally representative data specifically designed to estimate the prevalence of hypertension (HTN) and diabetes mellitus (DM) through standardized biomarker protocols.

### Sampling Technique

Both the 2017-18 and 2022 BDHS employed a consistent two-stage stratified cluster sampling design. The sampling frames were based on Enumeration Areas (EAs) defined by the Bangladesh Bureau of Statistics (BBS), with each EA, the Primary Sampling Unit (PSU), comprising approximately 120 households.

- First Stage: In both survey rounds, 675 PSUs were selected with probability proportional to the EA size. For the 2022 survey, this included 237 urban and 438 rural PSUs.
- Second Stage: The sampling intensity at the household level increased over time. While the 2017-18 survey selected 30 households per PSU, the 2022 survey selected an average of 45 households per PSU, resulting in a larger total sample of approximately 30,375 households in the most recent round.

### Biomarker Collection and Sub-sampling

Detailed information on DM and HTN was derived from the Biomarker Questionnaire. In both surveys, biomarker data, including fasting blood glucose and blood pressure measurements, were collected from a subsample of households (one-fourth of the selected households in 2017-18 and a designated subsample in 2022). Eligible participants included men and women aged 18 years and older residing in these households.

### Outcome Measures

This study encompasses two primary dependent variables: the presence or absence of diabetes and the presence or absence of hypertension. Individuals were classified as having elevated blood glucose or diabetes if their fasting blood glucose (FBG) exceeded a threshold of 6 mmol/L (1, 2). Otherwise, they were regarded to be in a normal health status or non-diabetic. Participants were categorized as having hypertension if, during the survey, their mean systolic blood pressure (SBP) exceeded 140 mmHg or their mean diastolic blood pressure (DBP) exceeded 90 mmHg. The categorization process involved adhering to the recommendations provided by the World Health Organization (WHO) and conducting a comprehensive evaluation of relevant literature.

### Explanatory Variables

The selection of explanatory variables is informed by a comprehensive evaluation of relevant literature. These variables are further categorized into demographic, biomedical, and behavioral aspects.

The demographic factors considered in this study include age (categorized as <=40 years and >40 years), gender (classified as female and male), division of residence (Barisal, Chittagong, Dhaka, Khulna, Mymensingh, Rajshahi, Rangpur, Sylhet), type of residence (categorized as rural and urban), education level (ranging from no education to primary, secondary, and higher education), and wealth index (categorized as poorest, poorer, middle, richer, and richest).

Biomedical variables encompass body mass index (BMI) categories, including thin (BMI < 18.5), normal (BMI 18.5-24.9), and overweight (BMI > 24.9), as well as the presence of hypertension and diabetes. Diabetes is recognized as a contributing factor to the development of hypertension, while hypertension is similarly acknowledged as a contributing factor to the development of diabetes. The behavioral characteristics under consideration include smoking status (yes/no).

### Statistical Analysis

All data management and statistical computations were executed using Stata (Version 17.0) and R (Version 4.5.1). To ensure the results provided nationally representative estimates, we applied sampling weights to adjust for the complex two-stage stratified cluster sampling design and participant non-response. The study population’s baseline characteristics were summarized using descriptive statistics, specifically frequency distributions and percentages for all sociodemographic and lifestyle covariates.

To evaluate initial associations between these covariates and the prevalence of Diabetes and Hypertension, we conducted bivariate analyses using contingency tables. Pearson’s Chi-square test was employed to assess statistical significance, strictly adhering to the validity assumption that all expected cell frequencies exceeded five.

Acknowledging the hierarchical structure inherent in the survey design, where observations within the same cluster are correlated, we adopted a mixed-effect logistic regression framework rather than a standard single-level model. This robust approach allows for the simultaneous estimation of individual-level risk factors while controlling for unobserved heterogeneity at the cluster level. Furthermore, the Intracluster Correlation Coefficient (ICC) was calculated to quantify the proportion of total variance attributable to community-level clustering, thereby validating the statistical necessity of the multilevel modeling approach.

To capture the full spectrum of cluster-level variability, the authors utilized generalized linear mixed models (GLMMs). The adjusted odds ratios (AORs) of the covariates were obtained from the mixed-effect logistic regression model. Let, *y*_*lP*_ be *p*^*th*^ individual/household from *l*^*th*^ cluster where *l* = 1,2,…,*q* and *p* = 1,2,…,*n*_*l*_; let, *x*_*lP*_ be a vector of covariates for *p*^*th*^ household from *l*^*th*^ cluster related with fixed effect parameter *β*. *u* represents a (*q* × 1) vector of random effects corresponding to *q* clusters and z_*lP*_ is a special vector of size (*q* × 1) which contains all zeros but a 1 at the *l*^*th*^ position; *l* = 1,2,…,*q*. Here, *μ*_*lp*_ = *E* (*Y*_*lp*_|*μ*_*l*_), where *μ*_*l*_ be the random effect of cluster *l*. Under GLMM the linear predictor takes the form, 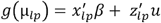. Using the logit link function in GLMM (61),

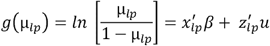

In this study, only random intercept has been considered, 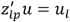, where, 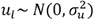. Therefore, the general conditional probability of *p*^*th*^ household from *l*^*th*^ cluster given the value of the explanatory variable of that observation and the random effect of that cluster is,

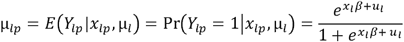

The likelihood function for the individuals corresponding to *l*^*th*^ cluster is,

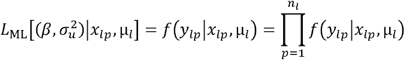

The estimates are obtained by maximizing the following marginal likelihood function,

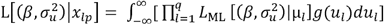

The formula for ICC is (62), 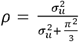.

Given the extensive number of covariates and the comparative nature of the study, relying solely on tabular data can obscure significant trends. Therefore, we utilized forest plots to synthesize the findings from the mixed-effect models. Although commonly used in meta-analysis, this approach is particularly valuable here for visualizing high-dimensional data. It allows readers to intuitively compare the effect sizes across the two survey waves and easily identify temporal shifts in risk factors, such as variables that gained or lost statistical significance between 2017-18 and 2022.

## Results

### Univariate Analysis

We initiated the analysis by examining the distribution of selected variables across the two survey periods, as detailed in Supplementary Table S1. The pooled analytic sample included 12,091 weighted participants from BDHS 2017-18 and 13,594 from BDHS 2022. In 2017-18, females constituted 57% of the sample, slightly decreasing to 55% in 2022. The age distribution shifted towards an older demographic over time; the proportion of participants aged over 40 years increased from 40% in 2017-18 to 44% in 2022. A notable shift was observed in body mass index (BMI) categories; the prevalence of overweight individuals decreased from 72% in 2017-18 to 64% in 2022, while the thin category rose from 4.7% to 7.2%. Regarding socioeconomic factors, the distribution of wealth remained relatively stable, with each quintile representing approximately 20% of the population in both surveys. Smoking prevalence dropped from 14% in 2017-18 to 10% in 2022.

The two survey waves reveal an unprecedented divergence in cardiometabolic disease burden (Figure 1). HTN prevalence declined modestly from 22% in 2017–18 to 15% in 2022; in stark contrast, DM prevalence increased dramatically, more than doubling from 23% to 49% within a four-year interval. This magnitude of change signals a rapid and substantial restructuring of metabolic risk in the Bangladeshi adult population.

**Figure 1:**
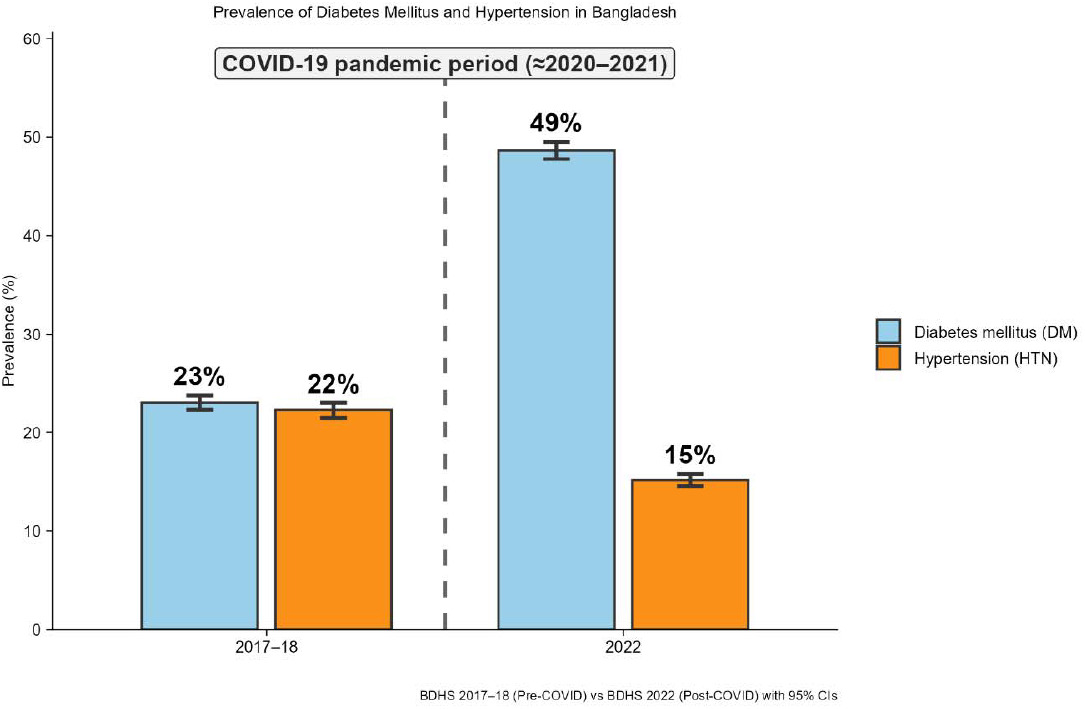
Prevalence shifts in diabetes mellitus and hypertension in Bangladesh (BDHS 2017–18 vs 2022).

### Bivariate analysis

#### Hypertension

The bivariate distribution of hypertension (Table 1) reveals distinct patterns. In both surveys, Age, BMI, Diabetes status, Division, Education, and Wealth Index were significantly associated with hypertension (p<0.001). Older age (>40 years) consistently showed higher hypertension rates (67% of cases in 2017-18 and 74% of cases in 2022). A divergence was noted in other factors. Sex was not associated with hypertension in 2017-18 (p=0.517) but became significant in 2022 (p<0.001), with females comprising a larger share of hypertensive cases. Similarly, Smoking Status was significantly associated with hypertension in 2017-18 (p=0.001) but lost significance in 2022 (p=0.951). Place of Residence (Urban/Rural) was not significantly associated with hypertension in either year.

**Table 1.**
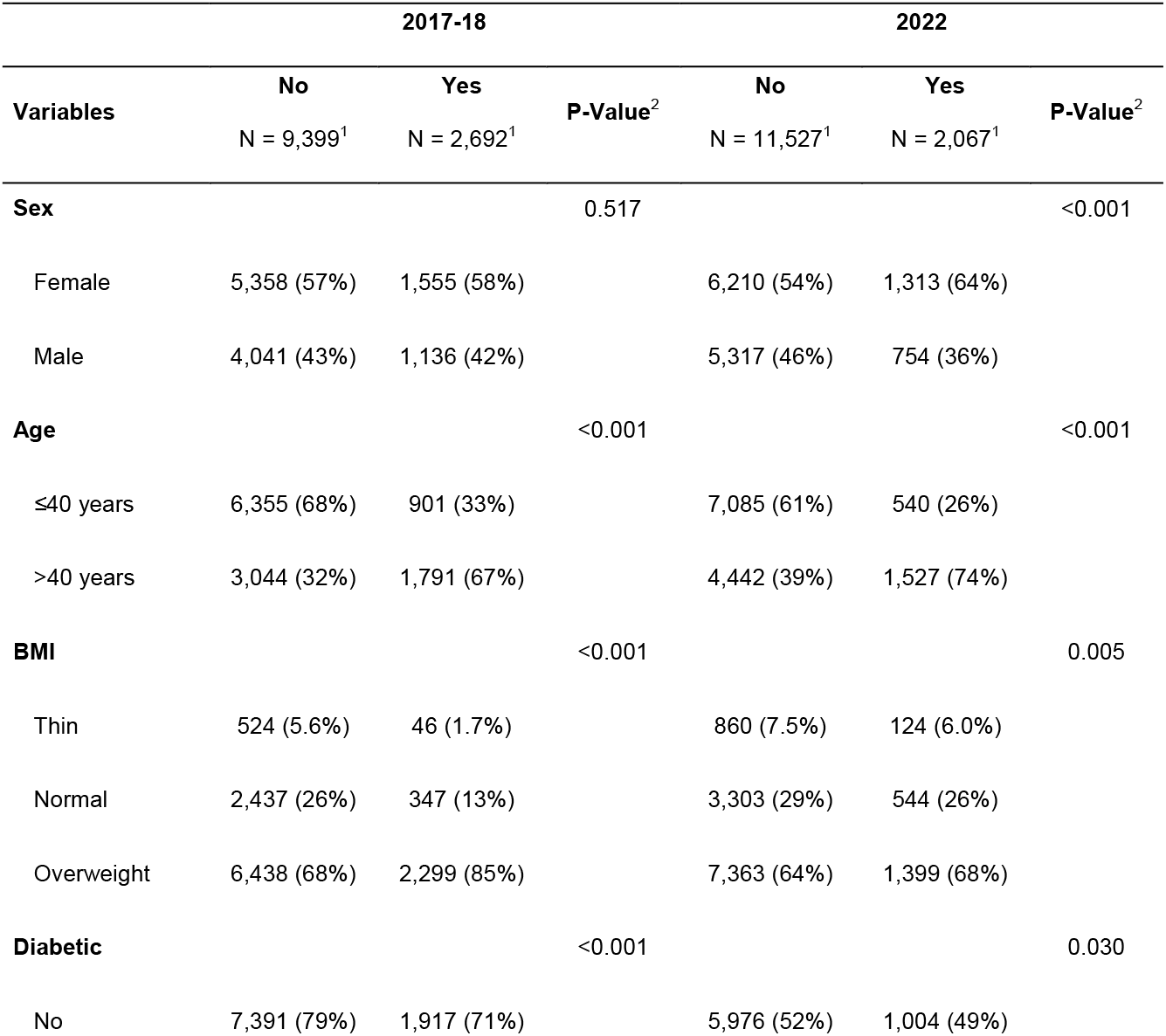
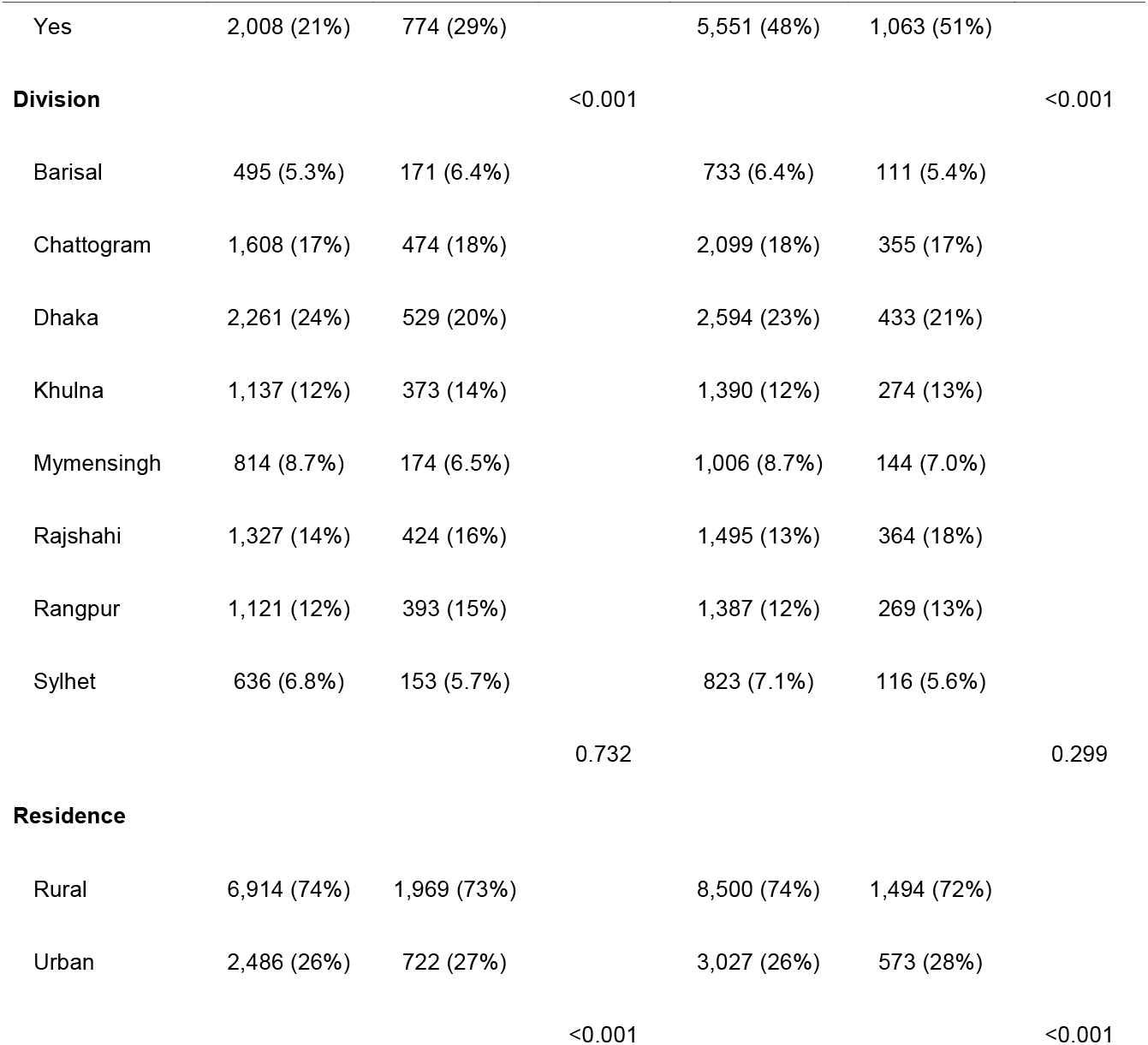
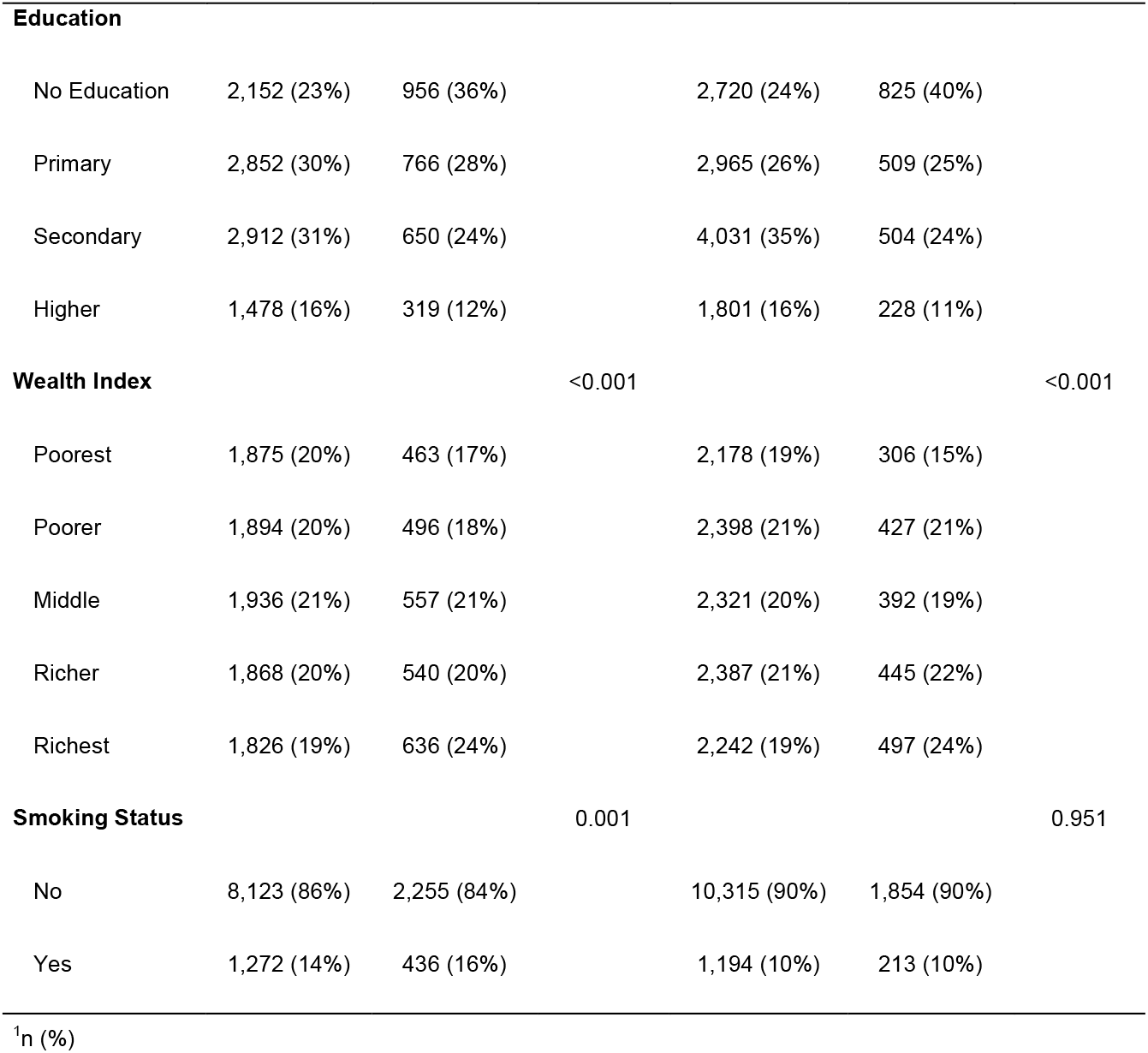
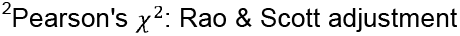
The distribution of Hypertension by sociodemographic variables.

#### Diabetes

The chi-square test was employed to assess the association between diabetes and selected covariates (Table 2). In both 2017-18 and 2022, Age group, BMI category, Hypertension status, administrative Division, place of Residence, and Wealth index were significantly associated with diabetes (p<0.001). In both years, individuals aged >40 years had a higher prevalence of diabetes compared to younger participants. The relationship with wealth was consistent; individuals in the Richest quintile exhibited the highest prevalence of diabetes in both 2017-18 (32%) and 2022 (24%). However, some associations evolved over time. Smoking status was not significantly associated with diabetes in 2017-18 (p=0.980) but showed a significant association in 2022 (p<0.001). Conversely, Education level was not statistically associated with diabetes in either survey year (p>0.05)

**Table 2.** The distribution of Diabetes by sociodemographic variables.

| Variables | 2017-18 |  | P-Value<sup>2</sup> | 2022 |  | P-Value<sup>2</sup> |
| --- | --- | --- | --- | --- | --- | --- |
|  | No<br>N = 9,308<sup>1</sup> | Yes<br>N = 2,783<sup>1</sup> |  | No<br>N = 6,980<sup>1</sup> | Yes<br>N = 6,614<sup>1</sup> |  |
| **sex** |  |  | 0.564 |  |  | 0.166 |
| Female | 5,336 (57%) | 1,577 (57%) |  | 3,903 (56%) | 3,620 (55%) |  |
| Male | 3,972 (43%) | 1,205 (43%) |  | 3,077 (44%) | 2,994 (45%) |  |
| **Age** |  |  | <0.001 |  |  | <0.001 |
| ≤40 years | 5,818 (63%) | 1,438 (52%) |  | 4,287 (61%) | 3,338 (50%) |  |
| >40 years | 3,490 (37%) | 1,345 (48%) |  | 2,692 (39%) | 3,277 (50%) |  |
| **BMI** |  |  | <0.001 |  |  | <0.001 |
| Thin | 474 (5.1%) | 96 (3.4%) |  | 579 (8.3%) | 405 (6.1%) |  |
| Normal | 2,265 (24%) | 518 (19%) |  | 2,108 (30%) | 1,739 (26%) |  |
| Overweight | 6,568 (71%) | 2,169 (78%) |  | 4,292 (61%) | 4,470 (68%) |  |
|  | No<br>N = 9,308<sup>1</sup> | Yes<br>N = 2,783<sup>1</sup> |  | No<br>N = 6,980<sup>1</sup> | Yes<br>N = 6,614<sup>1</sup> |  |
| **Hypertensive** |  |  | <0.001 |  |  | 0.030 |
| No | 7,391 (79%) | 2,008 (72%) |  | 5,976 (86%) | 5,551 (84%) |  |
| Yes | 1,917 (21%) | 774 (28%) |  | 1,004 (14%) | 1,063 (16%) |  |
| **Division** |  |  | <0.001 |  |  | <0.001 |
| Barisal | 504 (5.4%) | 163 (5.9%) |  | 459 (6.6%) | 385 (5.8%) |  |
| Chattogram | 1,575 (17%) | 506 (18%) |  | 1,143 (16%) | 1,312 (20%) |  |
| Dhaka | 1,805 (19%) | 986 (35%) |  | 1,303 (19%) | 1,724 (26%) |  |
| Khulna | 1,239 (13%) | 271 (9.7%) |  | 944 (14%) | 720 (11%) |  |
| Mymensingh | 800 (8.6%) | 188 (6.7%) |  | 702 (10%) | 449 (6.8%) |  |
| Rajshahi | 1,458 (16%) | 293 (11%) |  | 1,154 (17%) | 706 (11%) |  |
| Rangpur | 1,302 (14%) | 212 (7.6%) |  | 825 (12%) | 831 (13%) |  |
| Sylhet | 625 (6.7%) | 164 (5.9%) |  | 452 (6.5%) | 488 (7.4%) |  |
| **Residence** |  |  | <0.001 |  |  | <0.001 |
| Rural | 7,050 (76%) | 1,833 (66%) |  | 5,426 (78%) | 4,568 (69%) |  |
| Urban | 2,259 (24%) | 949 (34%) |  | 1,554 (22%) | 2,047 (31%) |  |
|  | No<br>N = 9,308<sup>1</sup> | Yes<br>N = 2,783<sup>1</sup> |  | No<br>N = 6,980<sup>1</sup> | Yes<br>N = 6,614<sup>1</sup> |  |
| **Education** |  |  | 0.917 |  |  | 0.126 |
| No Education | 2,379 (26%) | 729 (26%) |  | 1,784 (26%) | 1,761 (27%) |  |
| Primary | 2,802 (30%) | 816 (29%) |  | 1,739 (25%) | 1,736 (26%) |  |
| Secondary | 2,739 (29%) | 823 (30%) |  | 2,372 (34%) | 2,164 (33%) |  |
| Higher | 1,385 (15%) | 411 (15%) |  | 1,082 (16%) | 947 (14%) |  |
| **Wealth Index** |  |  | <0.001 |  |  | <0.001 |
| Poorest | 1,943 (21%) | 395 (14%) |  | 1,461 (21%) | 1,023 (15%) |  |
| Poorer | 2,012 (22%) | 377 (14%) |  | 1,580 (23%) | 1,245 (19%) |  |
| Middle | 2,026 (22%) | 467 (17%) |  | 1,390 (20%) | 1,323 (20%) |  |
| Richer | 1,767 (19%) | 640 (23%) |  | 1,382 (20%) | 1,450 (22%) |  |
| Richest | 1,559 (17%) | 903 (32%) |  | 1,167 (17%) | 1,573 (24%) |  |
| **Smoking Status** |  |  | 0.980 |  |  | <0.001 |
| No | 7,991 (86%) | 2,387 (86%) |  | 6,409 (92%) | 5,760 (87%) |  |
| Yes | 1,315 (14%) | 393 (14%) |  | 563 (8.1%) | 844 (13%) |  |

| Variables | 2017-18 |  |  | 2022 |  |  |
| --- | --- | --- | --- | --- | --- | --- |
|  | No<br>N = 9,308<sup>1</sup> | Yes<br>N = 2,783<sup>1</sup> | P-Value<sup>2</sup> | No<br>N = 6,980<sup>1</sup> | Yes<br>N = 6,614<sup>1</sup> | P-Value<sup>2</sup> |

<sup>1</sup>n (%)
<sup>2</sup>Pearson's $\chi^2$ : Rao & Scott adjustment

### Multivariate Analysis

#### Hypertension

To account for potential confounders, we performed a mixed-effects logistic regression (Table 3). Age was the strongest predictor in both years, with the odds of hypertension for those aged >40 years increasing from 3.57 (95% CI: 3.20-3.98) in 2017-18 to 4.03 (95% CI: 3.57-4.56) in 2022. The protective effect of Education was consistent; higher education was associated with lower odds of hypertension in both 2017-18 (AOR: 0.72) and 2022 (AOR: 0.64) compared to no education. Wealth also displayed a consistent graded association; the richest individuals had significantly higher odds of hypertension in both 2017-18 (AOR: 1.66) and 2022 (AOR: 1.78). BMI showed variation: being overweight was a significant risk factor in 2017-18 (AOR: 1.67) but was not significant in the 2022 mixed model (AOR: 1.06; 95% CI: 0.94-1.19). Similarly, Diabetes status was a significant predictor in 2017-18 (AOR 1.33) but was not significant in 2022 (p=0.594).

**Table 3.** Adjusted odds ratio (AOR), 95% confidence intervals, and P-value from mixed effect logistic regression model for hypertension.

| Variables | Adjusted<br>OR | 95% CI | p-value | Adjusted<br>OR | 95% CI | p-value |
| --- | --- | --- | --- | --- | --- | --- |
| Age |  |  | <0.001 |  |  | <0.001 |
| ≤40 years | — | — |  | — | — |  |
| >40 years | 3.57 | 3.20, 3.98 |  | 4.03 | 3.57, 4.56 |  |
| BMI |  |  | <0.001 |  |  | 0.019 |
| Normal | — | — |  | — | — |  |
| Thin | 0.61 | 0.44, 0.85 |  | 0.79 | 0.63, 0.98 |  |
| Overweight | 1.67 | 1.46, 1.90 |  | 1.06 | 0.94, 1.19 |  |
| Diabetic |  |  | <0.001 |  |  | 0.594 |
| No | — | — |  | — | — |  |
| Yes | 1.33 | 1.19, 1.48 |  | 1.03 | 0.92, 1.15 |  |
| Division |  |  | <0.001 |  |  | 0.003 |
| Barisal | — | — |  | — | — |  |
| Chattogram | 0.77 | 0.60, 1.00 |  | 1.08 | 0.81, 1.44 |  |
| Dhaka | 0.57 | 0.44, 0.74 |  | 0.99 | 0.74, 1.31 |  |
| Khulna | 0.83 | 0.64, 1.08 |  | 1.14 | 0.85, 1.54 |  |
| Mymensingh | 0.58 | 0.43, 0.78 |  | 0.89 | 0.65, 1.23 |  |
| Rajshahi | 0.91 | 0.70, 1.18 |  | 1.44 | 1.08, 1.93 |  |
| Rangpur | 1.06 | 0.82, 1.38 |  | 1.27 | 0.94, 1.70 |  |
| Sylhet | 0.66 | 0.49, 0.89 |  | 0.93 | 0.66, 1.30 |  |
| Education |  |  | 0.005 |  |  | <0.001 |
| No Education | — | — |  | — | — |  |
| Primary | 0.87 | 0.77, 0.99 |  | 0.80 | 0.70, 0.92 |  |
| Secondary | 0.86 | 0.74, 0.99 |  | 0.71 | 0.62, 0.83 |  |
| Higher | 0.72 | 0.60, 0.87 |  | 0.64 | 0.53, 0.78 |  |
| Wealth Index |  |  | <0.001 |  |  | <0.001 |
| Poorest | — | — |  | — | — |  |
| Poorer | 1.06 | 0.90, 1.24 |  | 1.24 | 1.05, 1.48 |  |
| Middle | 1.20 | 1.03, 1.41 |  | 1.24 | 1.04, 1.49 |  |
| Richer | 1.32 | 1.12, 1.56 |  | 1.45 | 1.21, 1.74 |  |
| Richest | 1.66 | 1.39, 1.99 |  | 1.78 | 1.47, 2.17 |  |
| Smoking Status |  |  | 0.007 |  |  | 0.846 |
| No | — | — |  | — | — |  |
| Yes | 0.83 | 0.73, 0.95 |  | 1.02 | 0.84, 1.23 |  |

#### Diabetes

The mixed-effects logistic regression for hypertension (Table 4) highlighted sustained risk factors. Age remained a robust predictor in both periods. Individuals aged > 40 years had significantly higher odds of diabetes in both 2017-18 (AOR: 1.66; 95% CI: 1.50-1.83) and 2022 (AOR: 1.89; 95% CI: 1.74-2.05) compared to those aged ≤ 40 years. The impact of BMI shifted; being overweight was not statistically significant in 2017-18 (AOR 1.11; p=0.144) but became a significant risk factor in 2022 (AOR: 1.22; 95% CI: 1.11-1.33). Hypertension status was consistently associated with higher odds of diabetes in both 2017-18 (AOR: 1.35) and 2022 (AOR: 1.17). Notably, the urban-rural divide widened. Place of residence was not significant in 2017-18 (AOR: 1.06; p=0.492) but in 2022, urban residents had 62% higher odds of diabetes compared to rural residents (AOR: 1.62; 95% CI: 1.31-2.01). Wealth status maintained a positive gradient, though the magnitude attenuated slightly; the richest quintile had 2.05 times higher odds in 2017-18 and 1.39 times higher odds in 2022 compared to the poorest.

**Table 4.** Adjusted odds ratio (AOR), 95% confidence intervals, and P-value from mixed effect logistic regression model for diabetes.

| Variables | Adjusted<br>OR | 95% CI | p-value | Adjusted<br>OR | 95% CI | p-value |
| --- | --- | --- | --- | --- | --- | --- |
| Age |  |  | <0.001 |  |  | <0.001 |
| ≤40 years | — | — |  | — | — |  |
| >40 years | 1.66 | 1.50, 1.83 |  | 1.89 | 1.74, 2.05 |  |
| BMI |  |  | 0.144 |  |  | <0.001 |
| Normal | — | — |  | — | — |  |
| Thin | 0.96 | 0.74, 1.25 |  | 0.83 | 0.70, 0.98 |  |
| Overweight | 1.11 | 0.99, 1.26 |  | 1.22 | 1.11, 1.33 |  |
| Hypertensive |  |  | <0.001 |  |  | 0.009 |
| No | — | — |  | — | — |  |
| Yes | 1.35 | 1.21, 1.52 |  | 1.17 | 1.04, 1.31 |  |
| Division |  |  | <0.001 |  |  | <0.001 |
| Barisal | — | — |  | — | — |  |
| Chattogram | 0.85 | 0.63, 1.15 |  | 1.50 | 1.00, 2.25 |  |
| Dhaka | 1.51 | 1.12, 2.03 |  | 1.70 | 1.14, 2.54 |  |
| Khulna | 0.57 | 0.41, 0.78 |  | 0.88 | 0.58, 1.34 |  |
| Mymensingh | 0.72 | 0.51, 1.00 |  | 0.78 | 0.51, 1.20 |  |
| Rajshahi | 0.56 | 0.41, 0.77 |  | 0.70 | 0.46, 1.06 |  |
| Rangpur | 0.49 | 0.35, 0.67 |  | 1.27 | 0.83, 1.92 |  |
| Sylhet | 0.72 | 0.51, 1.02 |  | 1.49 | 0.96, 2.32 |  |
| Residence |  |  | 0.492 |  |  | <0.001 |
| Rural | — | — |  | — | — |  |
| Urban | 1.06 | 0.90, 1.25 |  | 1.62 | 1.31, 2.01 |  |
| Wealth Index |  |  | <0.001 |  |  | <0.001 |
| Poorest | — | — |  | — | — |  |
| Poorer | 0.87 | 0.73, 1.04 |  | 1.00 | 0.87, 1.14 |  |
| Middle | 1.04 | 0.87, 1.23 |  | 1.02 | 0.89, 1.18 |  |
| Richer | 1.40 | 1.18, 1.67 |  | 1.25 | 1.08, 1.44 |  |
| Richest | 2.05 | 1.71, 2.46 |  | 1.39 | 1.19, 1.62 |  |

#### Forest Plot

Figure 2 presents the forest plot of adjusted odds ratios (AOR) for the determinants of diabetes and hypertension, visually comparing the magnitude of associations between the 2017-18 and 2022 survey periods.

**Figure 2:**
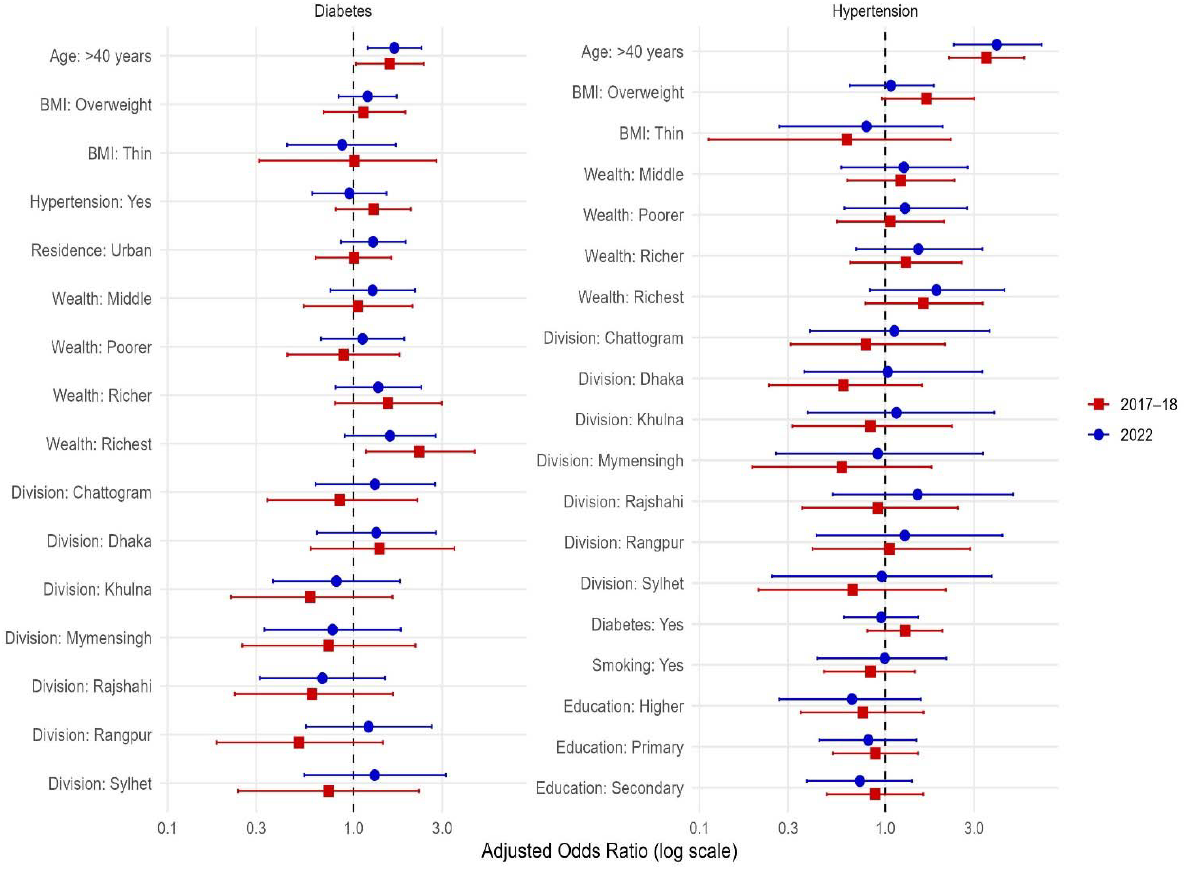
Comparative forest plot of adjusted odds ratios for determinants of Diabetes and Hypertension (BDHS 2017-18 vs. 2022)

The left panel of Figure 2 illustrates the factors associated with Diabetes Mellitus. The plot emphasizes the emergence of urban residence as a significant determinant in the post-pandemic period; in 2022, urban residents exhibited 62% higher odds of having diabetes (AOR: 1.62; 95% CI: 1.31-2.01) compared to rural residents, a significant increase from the non-significant association observed in 2017-18 (AOR: 1.06). Age remained a consistent predictor, with individuals aged over 40 years demonstrating 89% greater odds (AOR: 1.89; 95% CI: 1.74-2.05) in 2022 compared to those aged 40 years or younger. The wealth gradient persisted but showed attenuation; individuals in the richest wealth quintile had 105% higher odds (AOR: 2.05) in 2017-18, which decreased to 39% higher odds (AOR: 1.39; 95% CI: 1.19-1.62) in 2022 relative to the poorest quintile. Additionally, hypertensive individuals showed 17% higher odds of diabetes (AOR: 1.17) in 2022.

The right panel of Figure 2 displays the odds ratios for Hypertension. Age was the most dominant driver, with the odds of hypertension for individuals aged over 40 years increasing from 3.57 in 2017-18 to 4.03 times (AOR: 4.03; 95% CI: 3.57-4.56) in 2022 compared to the younger age group. A marked shift was observed regarding body mass index; while overweight individuals had 67% higher odds (AOR: 1.67) in 2017-18, this association was not statistically significant in 2022 (AOR: 1.06; 95% CI: 0.94-1.19). Education demonstrated a consistent protective effect; in 2022, individuals with secondary and higher education exhibited 29% (AOR: 0.71) and 36% (AOR: 0.64) lower odds of hypertension, respectively, compared to those with no education. Conversely, the wealthiest individuals experienced 78% higher odds (AOR: 1.78; 95% CI: 1.47-2.17) of hypertension in 2022 compared to the poorest group.

#### Model Selection

To identify the most appropriate statistical models, both fixed-effect and mixed-effects logistic regression models were fitted for all outcomes. The mixed-effects logistic regression models, incorporating a random intercept at the Primary Sampling Unit (PSU) level, were selected for the final interpretation to effectively account for the hierarchical clustering of the survey design. Model performance comparisons, detailed in Supplementary Table S4, confirmed that the mixed-effects approach provided a superior fit, evidenced by consistently lower Akaike Information Criterion (AIC) values across both survey years. For instance, in the 2022 diabetes model, the AIC demonstrated a substantial reduction from 18,222.1 in the fixed-effect model to 16,332.6 in the mixed-effects model, indicating a significant improvement in model fit. This superiority was statistically validated by Likelihood Ratio Tests (LRTs), which yielded p-values <0.001 for both hypertension and diabetes across both survey rounds.

Furthermore, the ICC underscored the necessity of accounting for cluster-level heterogeneity. The ICC values for hypertension were 0.046 in 2017-18 and 0.061 in 2022, indicating that 4.6% to 6.1% of the variance in hypertension status was attributable to between-cluster differences. Notably, the clustering effect was more pronounced for diabetes, with the ICC rising from 0.111 in 2017-18 to 0.281 in 2022. Multicollinearity was assessed prior to modelling, and no problematic multicollinearity was observed (all Generalized Variance Inflation Factors < 5).

## Discussion

This study examined both changes in disease prevalence and shifts in associated risk factors for DM and HTN among Bangladeshi adults before and after the COVID-19 pandemic. Using two nationally representative BDHS datasets (2017–18 and 2022), we demonstrate a pronounced divergence in NCD trajectories: diabetes prevalence nearly doubled, while hypertension prevalence declined. Beyond documenting these temporal trends, the analysis identifies how demographic, socioeconomic, biomedical, and community-level determinants evolved in the post-pandemic period. By integrating prevalence patterns with multilevel risk factor analysis, this study provides a comprehensive assessment of how the pandemic reshaped the NCD landscape in Bangladesh.

### Key Findings and Contextualization

The most striking observation is the shift of this disease burden: diabetes prevalence more than doubled from 23% in 2017-18 to 49% in 2022. This abrupt rise aligns with a growing body of global evidence linking SARS-CoV-2 infection to metabolic dysregulation. Recent meta-analyses have reported a 46% to 60% increased risk of diabetes following COVID-19 infection, suggesting that the pandemic functioned as a mass accelerator for the disease (59, 61). This surge is likely driven by the direct viral damage to pancreatic beta-cells and the indirect effects of pandemic-induced lifestyle changes (63). In the context of Bangladesh, specific gut microbiome dysbiosis observed in post-COVID patients with T2DM may further explain this localized surge(64). In contradiction, our study observed a decline in hypertension prevalence (22% to 15%), a finding that diverges from longitudinal studies where hypertension incidence increased post-pandemic (65). This difference may be partially attributed to survival bias. Evidence from Bangladesh indicates that patients with co-occurring CVD (Cardiovascular Disease), T2DM, and COVID-19 faced a 3.65-times higher risk of fatality (66). It is plausible that a significant cohort of hypertensive individuals were lost to COVID-19 mortality, statistically suppressing the prevalence in the surviving 2022 population(67, 68).

The mixed-effect logistic regression models revealed an evolving relationship between DM and HTN, where the robust bidirectional association observed in 2017-18 partially dissociated in the post-pandemic period. While hypertensive individuals in 2022 continued to face significantly higher odds of developing diabetes (AOR = 1.17; 95% CI: 1.04-1.31), diabetes status lost its statistical significance as an independent predictor for hypertension (AOR = 1.03; p=0.594). This divergence, potentially reflecting improved NCD management or COVID-19 related survival bias, underscores the critical need for integrated screening programs as these conditions continue to intensify morbidity in Bangladesh (63–65).

Age emerged as a predominant predictor for both conditions. In 2022, individuals over 40 years not only faced 4.03 times higher odds of HTN but also exhibited a profoundly increased risk for DM (AOR = 1.89; 95% CI: 1.74–2.05). These parallel trends corroborate studies highlighting the rising incidence of cardiometabolic NCDs among aging populations, as cumulative exposure to lifestyle risk factors, progressive pancreatic beta-cell decline, and reduced metabolic resilience jointly drive disease progression (69). Furthermore, post-COVID cohort data indicate that older populations (≥40 years) endured the most severe acute viral outcomes, triggering systemic inflammation that may further exacerbate subsequent NCD complications and rapidly accelerate the clinical onset of diabetes (67, 70).

Body mass index (BMI) emerged as a central determinant of both DM and HTN. Overweight individuals exhibited significantly higher odds of DM in both survey periods; however, the influence of obesity became more pronounced in the post-pandemic period. This pattern aligns with findings from a national survey reporting a marked increase in obesity-related odds ratios for diabetes in 2022(71). Similar evidence from Bangladesh has demonstrated that a BMI ≥22.5 kg/m^2^ elevates the risk of both DM and HTN (72–74). Interestingly, overweight status was no longer significantly associated with hypertension in 2022, in contrast to earlier observations. Nevertheless, normal BMI remained significantly protective against elevated blood pressure when compared with obesity, consistent with recent regional findings (62).

Socioeconomic disparities reflected a dual burden of non-communicable diseases. Individuals from higher wealth quintiles had significantly greater odds of DM, consistent with studies linking affluence to calorie-dense diets and sedentary lifestyles (71). However, this socioeconomic gradient reduced in 2022, with adjusted odds ratios declining from 2.27 to 1.57, suggesting a possible democratization of diabetes risk. In contrast, lower SES remained significantly associated with HTN prevalence, likely driven by psychosocial stress, limited access to healthcare, and poorer dietary quality (75). Higher educational attainment continued to demonstrate a protective effect against HTN, plausibly mediated through improved health literacy and healthier behavioral practices. This observation is consistent with recent evidence indicating that the pandemic widened educational inequalities in cardiovascular risk (71).

Substantial geographic and community-level disparities were also evident. Residents of the Dhaka division continued to experience a disproportionately high burden of NCDs, potentially reflecting rapid urbanization and greater exposure to processed foods. A novel finding was the emergence of urban residence as a significant risk factor for diabetes in 2022 (AOR = 1.62), a relationship not observed in 2017-18. This result is consistent with regional studies showing that predominantly urban occupational groups, such as private and self-employed workers, faced heightened metabolic risks during the pandemic(62). Moreover, the ICC values in diabetes model increased prominently to 0.281 in 2022, indicating that 28.1% of the variability in diabetes prevalence was attributable to community-level factors. This sharp rise underscores the growing importance of contextual determinants, such as neighborhood healthcare capacity and local economic resilience, in shaping post-pandemic diabetes risk and reinforces the value of a mixed-effects analytical approach.

### Strengths and Limitations

Our study has several distinct strengths that enhance the reliability of these findings. Primarily, the use of nationally representative data from two critical time points, the 2017-18 and 2022 BDHS survey allow for a uniquely robust comparison of the epidemiological landscape before and after the COVID-19 pandemic. This ensures that our insights are generalizable not just to the adult population of Bangladesh but also offer a reference point for other LMICs navigating similar post-pandemic transitions. Furthermore, we avoided the pitfalls of self-reported health data by relying on objective biomarker measurements for blood glucose and blood pressure, strictly adhering to WHO guidelines to minimize misclassification bias. Methodologically, the application of mixed-effect logistic regression models was a crucial choice; by accounting for the hierarchical structure of the data, we were able to quantify the significant rise in community-level variance (ICC) for diabetes, a nuance that conventional regression models would have missed. Finally, by adjusting for a comprehensive selection of sociodemographic and behavioral covariates, we ensured a rigorous assessment of the shifting risk profiles.

We must, however, acknowledge several limitations. The cross-sectional nature of the surveys prevents us from establishing causal pathways; while we observe strong associations, we cannot definitively prove the temporal sequence of exposure and disease onset at the individual level. A specific challenge in this comparative analysis was the lack of direct data on participants’ COVID-19 infection history or vaccination status, limiting our ability to directly link viral exposure to metabolic outcomes. We also cannot rule out survival bias as a contributing factor to the observed decline in hypertension; it is plausible that individuals with severe comorbidities faced higher mortality during the pandemic, potentially suppressing prevalence rates in the 2022 dataset. Additionally, while biomarkers ensured accurate diagnosis, certain risk factors like smoking and media exposure relied on self-reporting, which is subject to recall bias. Finally, the absence of detailed data on dietary habits, physical activity, and genetic factors in the BDHS limits our ability to thoroughly examine the behavioral mechanisms driving these trends.

To bridge these gaps, future research needs to shift toward longitudinal or cohort studies that can distinguish between temporary pandemic shocks and permanent epidemiological changes. There is also a clear need for qualitative research to unpack the urban penalty we observed, alongside studies incorporating more detailed data on diet and physical activity to better understand the behavioral roots of this diabetes rise.

## Conclusion

The burden of NCDs in Bangladesh has shifted markedly in the aftermath of the COVID-19 pandemic. While HTN prevalence declined, DM prevalence nearly doubled within a four-year period, indicating a rapid escalation of metabolic risk. This contrast suggests that the pandemic did not uniformly affect NCDs but instead accelerated DM through a combination of biological effects, lifestyle disruption, and uneven access to care.

The analysis further shows that DM risk is increasingly shaped by where people live, not only by who they are. The emergence of urban residence and the sharp rise in community-level variability highlight the growing importance of local environments, health system capacity, and socioeconomic context. Additionally, our multilevel modeling proves that geographic context matters more than ever in the post-pandemic era. Furthermore, the weakening of the previously bidirectional relationship between diabetes and hypertension suggests changing survival patterns and post-pandemic treatment dynamics.

Achieving SDG Target 3.4 will require a strategic shift from uniform national approaches toward targeted, community-specific interventions, particularly in rapidly urbanizing areas. Strengthening early detection, improving continuity of care, and addressing urban metabolic risk factors are essential to prevent diabetes from becoming an overwhelming long-term public health burden in Bangladesh.

## List of abbreviations

NCDs: Non-Communicable Diseases
HTN: Hypertension
T2DM: Type 2 Diabetes Mellitus
BDHS: Bangladesh Demographic and Health Survey
AOR: Adjusted Odds Ratio
ICC: Intracluster Correlation Coefficient
LRT: Likelihood Ratio Test
NIPORT: National Institute of Population Research and Training
AIC: Akaike Information Criterion
SES: Socioeconomic Status

## Declarations

### Ethics Approval and consent to participate

No ethical approval was needed for this study because it was based on secondary analysis of the data obtained from the 2022 BDHS (Bangladesh Demographic and Health Survey). The data was completely anonymous, and this study uses data with no identifiable information on the survey participants.

### Consent for publication

Not applicable.

### Availability of Data and Materials

The data in this article comes from the DHS Program database. This data can be found here: [https://dhsprogram.com/data]. The datasets generated during and/or analyzed during the current study are available from the corresponding author upon reasonable request. The Code used during the current study is available from the corresponding author upon reasonable request. Please contact the corresponding author for further information.

### Competing interests

The authors declare no competing interests

### Funding

Not applicable.

### Authors’ Contributions

KSAN contributed to the conceptualization, design, data curation, formal analysis, data interpretation, and original draft. AH contributed to data curation, formal analysis, data interpretation, and manuscript reviewing and editing. TJ contributed to the conceptualization and manuscript reviewing. All authors read and approved of the final manuscript.

## Acknowledgement

The authors thank the DHS Program for granting access to the BDHS 2017-18 and 2022 data.

## Notes

### Competing Interest Statement

The authors have declared no competing interest.

### Funding Statement

This study did not receive any funding

### Author Declarations

No ethical approval was needed for this study because it was based on secondary analysis of the data obtained from the 2022 BDHS (Bangladesh Demographic and Health Survey). The data were completely anonymous and this study uses data with no identifiable information on the survey participants.

## References

1. World Health Organization. Noncommunicable diseases 2023 [February 7, 2026]. Available from: https://www.who.int/news-room/fact-sheets/detail/noncommunicable-diseases.

2. Organization WH. he top 10 causes of death 2020 [February 07, 2026]. Available from: https://www.who.int/news-room/fact-sheets/detail/the-top-10-causes-of-death.

3. Niessen LW, Mohan D, Akuoku JK, Mirelman AJ, Ahmed S, Koehlmoos TP, et al. Tackling socioeconomic inequalities and non-communicable diseases in low-income and middle-income countries under the Sustainable Development agenda. The Lancet. 2018;391(10134):2036–46.

4. Jan S, Laba T-L, Essue BM, Gheorghe A, Muhunthan J, Engelgau M, et al. Action to address the household economic burden of non-communicable diseases. The Lancet. 2018;391(10134):2047–58.

5. Sassi F, Belloni A, Mirelman AJ, Suhrcke M, Thomas A, Salti N, et al. Equity impacts of price policies to promote healthy behaviours. The Lancet. 2018;391(10134):2059–70.

6. Bertram MY, Sweeny K, Lauer JA, Chisholm D, Sheehan P, Rasmussen B, et al. Investing in non-communicable diseases: an estimation of the return on investment for prevention and treatment services. The Lancet. 2018;391(10134):2071–8.

7. Muka T, Imo D, Jaspers L, Colpani V, Chaker L, van der Lee SJ, et al. The global impact of non-communicable diseases on healthcare spending and national income: a systematic review. European journal of epidemiology. 2015;30(4):251–77.

8. WHO G. Global status report on noncommunicable diseases 2010. 2011.

9. Organization WH. Global monitoring report on financial protection in health 2021: World Health Organization; 2021.

10. World Health Organization. World Health Statistics. [Available from: https://www.who.int/health-topics/sustainable-development-goals.

11. Organization WH. Noncommunicable diseases 2019 [Available from: https://www.who.int/health-topics/noncommunicable-diseases.

12. Organization WH. Noncommunicable diseases country profiles 2018 Geneva 2018 [

13. Rahman MA, Halder HR, Yadav UN, Mistry SK. Prevalence of and factors associated with hypertension according to JNC 7 and ACC/AHA 2017 guidelines in Bangladesh. Scientific reports. 2021;11(1):15420.

14. Cho N, Shaw J, Karuranga S, Huang Y, da Rocha Fernandes J, Ohlrogge A. Diabetes research and clinical practice. Diabetes Res Clin Pract. 2018;138:271–81.

15. Pm K. Global burden of hypertension: analysis of worldwide data. Lancet. 2005;365:217–23.

16. Mills KT, Stefanescu A, He J. The global epidemiology of hypertension. Nature Reviews Nephrology. 2020;16(4):223–37.

17. Neupane D, McLachlan CS, Sharma R, Gyawali B, Khanal V, Mishra SR, et al. Prevalence of hypertension in member countries of South Asian Association for Regional Cooperation (SAARC): systematic review and meta-analysis. Medicine. 2014;93(13):e74.

18. Organization WH. A multicountry analysis of noncommunicable disease surveillance data. World Health Organization 2010 [

19. Kyrou I, Tsigos C, Mavrogianni C, Cardon G, Van Stappen V, Latomme J, et al. Sociodemographic and lifestyle-related risk factors for identifying vulnerable groups for type 2 diabetes: a narrative review with emphasis on data from Europe. BMC endocrine disorders. 2020;20(Suppl 1):134.

20. Mamdouh H, Alnakhi WK, Hussain HY, Ibrahim GM, Hussein A, Mahmoud I, et al. Prevalence and associated risk factors of hypertension and pre-hypertension among the adult population: findings from the Dubai Household Survey, 2019. BMC Cardiovascular Disorders. 2022;22(1):18.

21. Mitchell N, Catenacci V, Wyatt HR, Hill JO. Obesity: overview of an epidemic. The Psychiatric clinics of North America. 2011;34(4):717.

22. Marbaniang SP, Chungkham HS, Lhungdim H. A structured additive modeling of diabetes and hypertension in Northeast India. Plos one. 2022;17(1):e0262560.

23. Willi C, Bodenmann P, Ghali WA, Faris PD, Cornuz J. Active smoking and the risk of type 2 diabetes: a systematic review and meta-analysis. Jama. 2007;298(22):2654–64.

24. Luo J, Rossouw J, Tong E, Giovino GA, Lee CC, Chen C, et al. Smoking and diabetes: does the increased risk ever go away? American journal of epidemiology. 2013;178(6):937–45.

25. Ramezankhani A, Parizadeh D, Azizi F, Hadaegh F. Sex differences in the association between diabetes and hypertension and the risk of stroke: cohort of the Tehran Lipid and Glucose Study. Biol Sex Differ. 2022;13(1):10.

26. Bank W. World Bank Open Data [February 07, 2026]. Available from: https://data.worldbank.org/indicator/SH.DTH.NCOM.ZS?locations=BD.

27. World Bank. [February 07, 2026]. Available from: https://www.worldbank.org/en/country/bangladesh/overview?utm.

28. Akhtar S, Nasir JA, Sarwar A, Nasr N, Javed A, Majeed R, et al. Prevalence of diabetes and pre-diabetes in Bangladesh: a systematic review and meta-analysis. BMJ open. 2020;10(9):e036086.

29. Mohiuddin A. Diabetes fact: Bangladesh perspective. Community and Public Health Nursing. 2019;4(1):39.

30. Chowdhury MZI, Rahman M, Akter T, Akhter T, Ahmed A, Shovon MA, et al. Hypertension prevalence and its trend in Bangladesh: evidence from a systematic review and meta-analysis. Clinical hypertension. 2020;26(1):10.

31. Statistics BBo. Population & Housing Census-2011: Union Statistics Dhaka 2014 [

32. Siddiquee T, Bhowmik B, Karmaker RK, Chowdhury A, Mahtab H, Khan AA, et al. Association of general and central obesity with diabetes and prediabetes in rural Bangladeshi population. Diabetes & Metabolic Syndrome: Clinical Research & Reviews. 2015;9(4):247–51.

33. Chowdhury MA, Uddin MJ, Haque MR, Ibrahimou B. Hypertension among adults in Bangladesh: evidence from a national cross-sectional survey. BMC Cardiovasc Disord. 2016;16:22.

34. Chowdhury MA, Uddin MJ, Khan HM, Haque MR. Type 2 diabetes and its correlates among adults in Bangladesh: a population based study. BMC Public Health. 2015;15:1070.

35. Rawal LB, Biswas T, Khandker NN, Saha SR, Bidat Chowdhury MM, Khan ANS, et al. Non-communicable disease (NCD) risk factors and diabetes among adults living in slum areas of Dhaka, Bangladesh. PLoS One. 2017;12(10):e0184967.

36. Riaz BK, Islam MZ, Islam AS, Zaman M, Hossain MA, Rahman MM, et al. Risk factors for non-communicable diseases in Bangladesh: findings of the population-based cross-sectional national survey 2018. BMJ open. 2020;10(11):e041334.

37. Sarker AR, Khanam M. Socio-economic inequalities in diabetes and prediabetes among Bangladeshi adults. Diabetology international. 2022;13(2):421–35.

38. Organization WH. Time to deliver: Report of the WHO independent high-level commission on noncommunicable diseases Geneva 2018 [

39. Nugent R, Bertram MY, Jan S, Niessen LW, Sassi F, Jamison DT, et al. Investing in non-communicable disease prevention and management to advance the Sustainable Development Goals. The Lancet. 2018;391(10134):2029–35.

40. Abarca-Gómez L, Abdeen ZA, Hamid ZA, Abu-Rmeileh NM, Acosta-Cazares B, Acuin C, et al. Worldwide trends in body-mass index, underweight, overweight, and obesity from 1975 to 2016: a pooled analysis of 2416 population-based measurement studies in 128· 9 million children, adolescents, and adults. The lancet. 2017;390(10113):2627–42.

41. Gray N, Picone G, Sloan F, Yashkin A. The relationship between BMI and onset of diabetes mellitus and its complications. Southern medical journal. 2015;108(1):29.

42. Mita G, Ni Mhurchu C, Jull A. Effectiveness of social media in reducing risk factors for noncommunicable diseases: a systematic review and meta-analysis of randomized controlled trials. Nutr Rev. 2016;74(4):237–47.

43. Pearson-Stuttard J, Zhou B, Kontis V, Bentham J, Gunter MJ, Ezzati M. Retracted: Worldwide burden of cancer attributable to diabetes and high body-mass index: a comparative risk assessment. The Lancet Diabetes & Endocrinology. 2018;6(2):95–104.

44. Organization WH. Global report on diabetes: Executive summary Geneva 2016 [

45. Roy PK, Khan MHR, Akter T, Rahman MS. Exploring socio-demographic-and geographical-variations in prevalence of diabetes and hypertension in Bangladesh: Bayesian spatial analysis of national health survey data. Spatial and spatio-temporal epidemiology. 2019;29:71–83.

46. Ali N, Akram R, Sheikh N, Sarker AR, Sultana M. Sex-specific prevalence, inequality and associated predictors of hypertension, diabetes, and comorbidity among Bangladeshi adults: results from a nationwide cross-sectional demographic and health survey. BMJ Open. 2019;9(9):e029364.

47. Rahman MM, Gilmour S, Akter S, Abe SK, Saito E, Shibuya K. Prevalence and control of hypertension in Bangladesh: a multilevel analysis of a nationwide population-based survey. Journal of hypertension. 2015;33(3):465–72.

48. Elfarra RM. A stakeholder analysis of noncommunicable diseases’ multisectoral action plan in Bangladesh. WHO South-East Asia Journal of Public Health. 2021;10(1):37–46.

49. International NIoPRaTNMaAI. Bangladesh Demographic and Health Survey 2011 Dhaka, Bangladesh; Calverton, Maryland, USA 2013 [

50. ICF NIoPRaTN. Bangladesh Demographic and Health Survey 2022: Key indicators report Dhaka, Bangladesh; Rockville, Maryland, USA 2023 [

51. Islam SMS, Purnat TD, Phuong NTA, Mwingira U, Schacht K, Fröschl G. Non-Communicable Diseases (NCDs) in developing countries: a symposium report. Globalization and health. 2014;10(1):81.

52. Organization WH. Diabetes 2023 [Available from: https://www.who.int/news-room/fact-sheets/detail/diabetes.

53. Organization WH. Hypertension 2019 [Available from: https://www.who.int/health-topics/hypertension.

54. Lim SS, Vos T, Flaxman AD, Danaei G, Shibuya K, Adair-Rohani H, et al. A comparative risk assessment of burden of disease and injury attributable to 67 risk factors and risk factor clusters in 21 regions, 1990–2010:a systematic analysis for the Global Burden of Disease Study 2010. The lancet. 2012;380(9859):2224–60.

55. Rodríguez G, Elo I. Intra-class Correlation in Random-effects Models for Binary Data. The Stata Journal. 2003;3(1):32–46.

56. Wakefield J. Multi-level modelling, the ecologic fallacy, and hybrid study designs. Int J Epidemiol. 2009;38(2):330–6.

57. Akter F, Haq A, Godman B, Chowdhury K, Kumar S, Haque M, editors. Impact of lockdown measures on health outcomes of adults with Type 2 diabetes mellitus in Bangladesh. Healthcare; 2023: MDPI.

58. Yan Y, Yang Y, Wang F, Ren H, Zhang S, Shi X, et al. Clinical characteristics and outcomes of patients with severe covid-19 with diabetes. BMJ Open Diabetes Res Care. 2020;8(1).

59. Wong R, Lam E, Bramante CT, Johnson SG, Reusch J, Wilkins KJ, et al. Does COVID-19 Infection Increase the Risk of Diabetes? Current Evidence. Curr Diab Rep. 2023;23(8):207–16.

60. Sayeed MA, Mahtab H, Khanam PA, Latif ZA, Banu A, Khan AK. Prevalence of diabetes and impaired fasting glucose in urban population of Bangladesh. Bangladesh Med Res Counc Bull. 2007;33(1):1–12.

61. Zhou J, Wang Y, Xu R. Association of COVID-19 infection and the risk of new incident diabetes: a systematic review and meta-analysis. Front Endocrinol (Lausanne). 2024;15:1429848.

62. Munam AM, Hossain A. Prevalence of Diabetes Mellitus and Hypertension Among COVID-19 Patients: A Cross-Sectional Study Exploring Associations With Sociodemographic and Biological Factors in Rajshahi Division, Bangladesh. Health Sci Rep. 2025;8(8):e71183.

63. Choi JH, Kim KM, Song K, Seo GH. Risk for Newly Diagnosed Type 2 Diabetes Mellitus after COVID-19 among Korean Adults: A Nationwide Matched Cohort Study. Endocrinol Metab (Seoul). 2023;38(2):245–52.

64. Mannan A, Hoque MN, Noyon SH, Mehedi HMH, Foysal MJ, Salauddin A, et al. SARS-CoV-2 infection alters the gut microbiome in diabetes patients: A cross-sectional study from Bangladesh. J Med Virol. 2023;95(4):e28691.

65. Trimarco V, Izzo R, Pacella D, Trama U, Manzi MV, Lombardi A, et al. Incidence of new-onset hypertension before, during, and after the COVID-19 pandemic: a 7-year longitudinal cohort study in a large population. BMC Med. 2024;22(1):127.

66. Sharif N, Sharif N, Khan A, Halawani IF, Alzahrani FM, Alzahrani KJ, et al. Prevalence and impact of long COVID-19 among patients with diabetes and cardiovascular diseases in Bangladesh. Front Public Health. 2023;11:1222868.

67. Islam R, Ahmed S, Chakma SK, Mahmud T, Al Mamun A, Islam Z, et al. Smoking and pre-existing co-morbidities as risk factors for developing severity of COVID-19 infection: Evidence from a field hospital in a rural area of Bangladesh. PLoS One. 2023;18(12):e0295040.

68. Li C, Islam N, Gutierrez JP, Gutiérrez-Barreto SE, Castañeda Prado A, Moolenaar RL, et al. Associations of diabetes, hypertension and obesity with COVID-19 mortality: a systematic review and meta-analysis. BMJ Glob Health. 2023;8(12).

69. Kirkman MS, Briscoe VJ, Clark N, Florez H, Haas LB, Halter JB, et al. Diabetes in older adults. Diabetes Care. 2012;35(12):2650–64.

70. Xie Y, Al-Aly Z. Risks and burdens of incident diabetes in long COVID: a cohort study. Lancet Diabetes Endocrinol. 2022;10(5):311–21.

71. Chiavarini M, Dolcini J, Firmani G, Ponzio E, Barbadoro P. Prevalence of Diabetes, Hypertension, and Associated of Cardiovascular Diseases: A Comparative Pre- and Post-COVID Study. Diseases. 2024;12(12):329.

72. Nahin KSA, Jannatul T. Risk factors for non-communicable diseases among Bangladeshi adults: an application of generalised linear mixed model on multilevel demographic and health survey data. BMJ Open. 2025;15(3):e082952.

73. Organization WH. Noncommunicable diseases: Progress monitor 2022 Institution Geneva2022 [

74. Marquess JG. The elderly and diabetes: an age trend and an epidemic converging. Consult Pharm. 2008;23 Suppl B:5–11.

75. Ahmed A, Rahman M, Hasan R, Shima SA, Faruquee MH, Islam T, et al. Hypertension and associated risk factors in some selected rural areas of Bangladesh. International Journal of Research in Medical Sciences. 2017;2(3):925–31.

