## Supplemental Table for "Shifting Prevalence and Risk Factors of Non-communicable Diseases in Bangladesh: A Comparative Multilevel Analysis of Nationally Representative BDHS Data (2017–2022)"

1. Background characteristics of the study population

| Variable | Weighted n (%) 2017–18 N = 12,091[1] | 2022 N = 13,594[1] |
|---|---|---|
| **Hypertensive** | | |
| No | 9,399 (78%) | 11,527 (85%) |
| Yes | 2,692 (22%) | 2,067 (15%) |
| **Diabetic** | | |
| No | 9,308 (77%) | 6,980 (51%) |
| Yes | 2,783 (23%) | 6,614 (49%) |
| **Age** | | |
| ≤40 years | 7,256 (60%) | 7,625 (56%) |
| >40 years | 4,835 (40%) | 5,969 (44%) |
| **Sex** | | |
| Female | 6,914 (57%) | 7,523 (55%) |
| Male | 5,177 (43%) | 6,071 (45%) |
| **Division** | | |
| Barisal | 666 (5.5%) | 844 (6.2%) |
| Chattogram | 2,082 (17%) | 2,454 (18%) |
| Dhaka | 2,791 (23%) | 3,027 (22%) |
| Khulna | 1,510 (12%) | 1,664 (12%) |
| Mymensingh | 988 (8.2%) | 1,150 (8.5%) |
| Rajshahi | 1,751 (14%) | 1,859 (14%) |
| Rangpur | 1,514 (13%) | 1,656 (12%) |
| Sylhet | 789 (6.5%) | 939 (6.9%) |
| **Residence** | | |
| Rural | 8,883 (73%) | 9,993 (74%) |
| Urban | 3,208 (27%) | 3,600 (26%) |

| | Weighted n (%) | |
|---|---|---|
| **Variable** | **2017–18** N = 12,091[1] | **2022** N = 13,594[1] |
| **Education** | | |
| No Education | 3,108 (26%) | 3,545 (26%) |
| Primary | 3,618 (30%) | 3,474 (26%) |
| Secondary | 3,562 (29%) | 4,536 (33%) |
| Higher | 1,797 (15%) | 2,029 (15%) |
| **Wealth Index** | | |
| Poorest | 2,338 (19%) | 2,484 (18%) |
| Poorer | 2,389 (20%) | 2,825 (21%) |
| Middle | 2,494 (21%) | 2,713 (20%) |
| Richer | 2,408 (20%) | 2,832 (21%) |
| Richest | 2,462 (20%) | 2,739 (20%) |
| **BMI** | | |
| Thin | 570 (4.7%) | 984 (7.2%) |
| Normal | 2,783 (23%) | 3,847 (28%) |
| Overweight | 8,737 (72%) | 8,762 (64%) |
| **Smoking Status** | | |
| No | 10,378 (86%) | 12,169 (90%) |
| Yes | 1,708 (14%) | 1,407 (10%) |

[1]n (%)

2. Adjusted OR (AOR), 95% CI and p value from fixed effect logistic regression model for Hypertension

| | | 2017–18 | | | 2022 | |
|---|---|---|---|---|---|---|
| **Variables** | **Adjusted OR** | **95% CI** | **p-value** | **Adjusted OR** | **95% CI** | **p-value** |
| **Age** | | | <0.001 | | | <0.001 |
| ≤40 years | — | — | | — | — | |
| >40 years | 3.49 | 3.13, 3.89 | | 3.98 | 3.47, 4.55 | |
| **BMI** | | | <0.001 | | | 0.019 |
| Normal | — | — | | — | — | |
| Thin | 0.62 | 0.43, 0.89 | | 0.79 | 0.63, 1.00 | |
| Overweight | 1.66 | 1.44, 1.92 | | 1.07 | 0.95, 1.21 | |
| **Diabetic** | | | <0.001 | | | 0.400 |
| No | — | — | | — | — | |
| Yes | 1.28 | 1.14, 1.44 | | 0.95 | 0.84, 1.07 | |
| **Division** | | | <0.001 | | | 0.004 |
| Barisal | — | — | | — | — | |
| Chattogram | 0.79 | 0.62, 0.99 | | 1.12 | 0.89, 1.41 | |
| Dhaka | 0.59 | 0.47, 0.75 | | 1.03 | 0.82, 1.29 | |
| Khulna | 0.83 | 0.67, 1.04 | | 1.15 | 0.91, 1.45 | |
| Mymensingh | 0.58 | 0.46, 0.74 | | 0.91 | 0.71, 1.16 | |
| Rajshahi | 0.91 | 0.72, 1.16 | | 1.49 | 1.16, 1.92 | |
| Rangpur | 1.05 | 0.85, 1.31 | | 1.27 | 1.02, 1.59 | |
| Sylhet | 0.67 | 0.53, 0.85 | | 0.96 | 0.74, 1.23 | |
| **Education** | | | 0.023 | | | <0.001 |
| No Education | — | — | | — | — | |
| Primary | 0.89 | 0.77, 1.01 | | 0.81 | 0.70, 0.94 | |

| | 2017–18 | | | 2022 | | |
|---|---|---|---|---|---|---|
| **Variables** | **Adjusted OR** | **95% CI** | **p-value** | **Adjusted OR** | **95% CI** | **p-value** |
| Secondary | 0.88 | 0.75, 1.03 | | 0.73 | 0.62, 0.86 | |
| Higher | 0.76 | 0.63, 0.91 | | 0.66 | 0.54, 0.81 | |
| **Wealth Index** | | | <0.001 | | | <0.001 |
| Poorest | — | — | | — | — | |
| Poorer | 1.07 | 0.90, 1.26 | | 1.28 | 1.05, 1.55 | |
| Middle | 1.21 | 1.00, 1.45 | | 1.26 | 1.04, 1.53 | |
| Richer | 1.29 | 1.07, 1.55 | | 1.51 | 1.24, 1.82 | |
| Richest | 1.61 | 1.33, 1.95 | | 1.88 | 1.52, 2.34 | |
| **Smoking Status** | | | 0.008 | | | 0.946 |
| No | — | — | | — | — | |
| Yes | 0.83 | 0.73, 0.95 | | 0.99 | 0.80, 1.23 | |

Abbreviations: CI = Confidence Interval, OR = Odds Ratio

3. Adjusted OR (AOR), 95% CI and p value from fixed effect logistic regression model for Diabetes

| | 2017–18 | | | 2022 | | |
|---|---|---|---|---|---|---|
| **Variables** | **Adjusted OR** | **95% CI** | **p-value** | **Adjusted OR** | **95% CI** | **p-value** |
| **Age** | | | <0.001 | | | <0.001 |
| ≤40 years | — | — | | — | — | |
| >40 years | 1.57 | 1.41, 1.74 | | 1.66 | 1.53, 1.80 | |
| **BMI** | | | 0.138 | | | <0.001 |

| | | 2017–18 | | | 2022 | |
|---|---|---|---|---|---|---|
| **Variables** | **Adjusted OR** | **95% CI** | **p-value** | **Adjusted OR** | **95% CI** | **p-value** |
| Normal | — | — | | — | — | |
| Thin | 1.01 | 0.77, 1.32 | | 0.87 | 0.75, 1.01 | |
| Overweight | 1.13 | 1.00, 1.29 | | 1.19 | 1.10, 1.30 | |
| **Hypertensive** | | | <0.001 | | | 0.399 |
| No | — | — | | — | — | |
| Yes | 1.29 | 1.14, 1.45 | | 0.95 | 0.85, 1.07 | |
| **Division** | | | <0.001 | | | <0.001 |
| Barisal | — | — | | — | — | |
| Chattogram | 0.84 | 0.63, 1.14 | | 1.30 | 0.96, 1.78 | |
| Dhaka | 1.38 | 1.02, 1.88 | | 1.33 | 0.96, 1.84 | |
| Khulna | 0.59 | 0.44, 0.79 | | 0.81 | 0.59, 1.11 | |
| Mymensingh | 0.73 | 0.55, 0.98 | | 0.77 | 0.54, 1.10 | |
| Rajshahi | 0.60 | 0.43, 0.83 | | 0.68 | 0.47, 0.98 | |
| Rangpur | 0.51 | 0.37, 0.69 | | 1.21 | 0.84, 1.73 | |
| Sylhet | 0.74 | 0.53, 1.02 | | 1.30 | 0.95, 1.78 | |
| **Residence** | | | 0.931 | | | 0.014 |
| Rural | — | — | | — | — | |
| Urban | 1.01 | 0.85, 1.19 | | 1.28 | 1.05, 1.55 | |
| **Wealth Index** | | | <0.001 | | | <0.001 |
| Poorest | — | — | | — | — | |
| Poorer | 0.89 | 0.71, 1.10 | | 1.12 | 0.96, 1.31 | |
| Middle | 1.06 | 0.84, 1.33 | | 1.27 | 1.07, 1.51 | |
| Richer | 1.53 | 1.22, 1.91 | | 1.36 | 1.14, 1.62 | |
| Richest | 2.27 | 1.82, 2.83 | | 1.57 | 1.30, 1.90 | |

Abbreviations: CI = Confidence Interval, OR = Odds Ratio

4. Akaike Information Criterions (AICs), p-value of Likelihood Ratio Tests (LRTs) and Intra-class Correlation Coefficients (ICCs) for both fixed and mixed effect models

| Survey year | AIC (Fixed) | AIC (Mixed) | LRT p-value | ICC |
|---|---|---|---|---|
| **Hypertension** | | | | |
| 2017–18 | 11,607.0 | 11,549.8 | <0.001 | 0.046 |
| 2022 | 10,609.0 | 10,524.0 | <0.001 | 0.061 |
| **Diabetes** | | | | |
| 2017–18 | 12,286.0 | 11,949.0 | <0.001 | 0.111 |
| 2022 | 18,222.1 | 16,332.6 | <0.001 | 0.281 |
